# Individual calf muscle structure-function adaptations to 12 weeks of eccentric training measured with 3D ultrasonography and dynamometry

**DOI:** 10.1101/2025.08.27.25334587

**Authors:** C. Rivares Benitez, G. Weide, R. T. Jaspers, M. Sartori

**Affiliations:** Department of Biomechanical Engineering, University of Twente, Enschede, Netherlands; Laboratory for Myology, Department of Human Movement Sciences, Faculty of Behavioural and Movement Sciences, Vrije Universiteit Amsterdam, Amsterdam Movement Sciences, Amsterdam, The Netherlands

**Keywords:** Adaptive capacity, muscle plasticity, eccentric exercise, muscle remodelling

## Abstract

Understanding skeletal muscle adaptation is key to optimizing training and rehabilitation strategies, yet the causal links between training stimuli and muscle response remain unclear. This gap reflects the difficulty of observing multi-scale adaptations within the same muscle in vivo. Calf muscles, critical for propulsion and postural control, remain relatively under-studied, and long-term outcomes with intermediate stages of adaptation are rarely documented.

We investigated temporal and regional remodelling of the gastrocnemius medialis muscle during 12 weeks of eccentric training in six young, healthy adults. Participants trained on alternate days with progressive overload calf raise exercises. Muscle architecture and function were assessed at baseline, 6, and 12 weeks using 3D ultrasonography and dynamometry.

Group-level analysis revealed a 38% increase in peak plantarflexion torque at 90° (p < 0.01), while muscle volume, PCSA, fascicle length, and pennation angle showed no consistent changes. Individual response profiles varied: some participants showed longitudinal growth with longer fascicles and smaller pennation angles, others displayed radial growth with increased PCSA, while some exhibited minimal architectural change despite torque gains. Trends suggested shifts in fascicle length-angle torque relationships and altered tendon compliance in certain individuals.

By combining regional muscle morphology with functional outcomes over time, this study demonstrates the feasibility of tracking multi-scale adaptation in vivo. The pronounced inter-individual and region-specific variability highlights the need for tailored interventions that consider baseline architecture and regional strain patterns to optimize outcomes in training and rehabilitation.

## Introduction

Understanding how to steer neuromuscular adaptations is essential for improving both rehabilitation outcomes and athletic performance. In clinical populations, such as individuals with neuromuscular disorders, muscular dystrophies, or age-related muscle decline, restoring function requires targeted interventions that address specific changes in muscle morphology and force-generating capacity. Similarly, in athletic settings, enhancing speed, power, or movement efficiency depends on fine-tuning muscle architecture through tailored training. Despite growing interest in personalized training interventions and performance optimization, current strategies often rely on generalized protocols that do not account for individual differences in muscle baseline, function and structure as well as adaptive capacity (Noone et al., 2024). However, there remains a fundamental knowledge gap: the causal links between training stimuli and muscle response are not fully understood, largely because it is difficult to capture multi-scale and multi-modal adaptations within the same muscle in vivo. Here, muscle morphology (global and regional) was measured together with force-generating capacity over 12 weeks, contributing to addressing this gap.

In calf muscles, eccentric muscle training is known for its potential to stimulate muscle hypertrophy, increase force-generating capacity, and induce architectural remodelling (Kruse et al., 2021; Duclay et al., 2009; Bizet et al., 2025; Geremia et al., 2019). Several studies have shown positive effects after short- and mid-term interventions (e.g., Duclay et al., 2009; Bizet et al., 2025). However, long-term outcomes and the progression of adaptations at intermediate stages are under-documented (Geremia et al., 2019). Given their role in both propulsion and postural control, adaptations in the calf muscles have implications not only for performance enhancement but also for clinical rehabilitation.

Morphological adaptations to training occur unevenly along the length of a muscle, with eccentric training often inducing greater changes in distal regions, such as increased fascicle length or hypertrophy, compared to mid or proximal areas (Benford et al. 2020; Pincheira et al. 2022; Andrews et al. 2024; Simpson et al., 2017). This highlights the importance of analysing muscle structure at a regional level, as whole-muscle averages may obscure meaningful, functionally relevant changes. Even if whole-muscle changes do not directly link to function, regional adaptations might still be relevant. Different muscle regions can respond differently to loading (Simpson et al., 2017; Crouzier et al., 2018) and play distinct roles in movement or joint mechanics. Ignoring that could mean missing relevant changes. In the soleus and gastrocnemius muscles, however, regional adaptations remain largely underexplored, with only limited evidence suggesting non-uniform remodelling in response to eccentric training.

Despite their crucial role, the gastrocnemius and soleus are often underrepresented in sports performance research (Green et al., 2022). In clinical contexts, the soleus has been identified as a key muscle to monitor in heart failure, as its reduced size closely correlates with impaired aerobic capacity, highlighting its potential as a sentinel marker for functional decline and rehabilitation needs (Panizzolo et al., 2015 and 2016). In neuromuscular disorders such as cerebral palsy and stroke, the calf muscles often become shortened or spastic, limiting ankle dorsiflexion and impairing gait (Barber et al., 2011; Yakut et al., 2024; van den Noort et al., 2017). The altered architecture often contributes to reduced mobility and functional independence (Bjornson et al., 2007; Sahoo et al., 2017; Schenker et al., 2005; Zwier et al., 2010). Targeted interventions addressing calf muscle structure and function are therefore essential in rehabilitation strategies for these populations. However, it is still unclear how improvements in calf muscle architecture directly impact cardiovascular health or neuromuscular recovery. This lack of clarity further underscores the need for detailed data on how these muscles respond to eccentric loading over time.

The time course of neuromuscular adaptation involves distinct phases that differ in their underlying mechanisms. Early changes are mostly neural and involve increases in sarcomere load and length range over which sarcomeres operate, while structural changes like fascicle lengthening and PCSA increases typically appear after several weeks (Folland and Williams, 2007; Kruse et al., 2021; Rong et al., 2025). These adaptations do not occur uniformly, and measuring only at the start and end may miss important intermediate changes that help explain functional outcomes.

In this study, we examined muscle architecture, morphology, and function simultaneously, in detail over a multi-week eccentric training intervention. By combining established physiological assessments, such as ultrasonography and dynamometry, with advanced imaging techniques, 3D ultrasonography, we aimed to gain deeper insight into how human skeletal muscle adapts over time. Specifically, we quantified regional changes in the medial gastrocnemius at multiple time points during a 12-week training program, which to our knowledge is the first investigation to capture both the temporal and regional progression of architectural adaptation.

## Materials and methods

### Participants characteristics

Six healthy and physical active volunteers (3 males and 3 females) were enrolled in the study. The participants had a mean age of 25 ± 4.4 years, height of 178.41 ± 9.9 cm, weight of 75.25 ± 16.2 kg at baseline and 75.33 ± 15.09 at week 12, tibia length of 37.5 ± 2.5 cm. Inclusion criteria were: not participating in any structured strength training program beyond their usual sport or physical activity; absence of musculoskeletal injury in the lower limbs; and ability to complete all training sessions and experimental assessments.

### Study design

This study is part of a larger experiment focused on neuromuscular adaptations in the lower leg following a training intervention. Before the first measurement session, participants completed two familiarization sessions to get used to the testing equipment, measurement protocols, and exercise instructions. These sessions were also used to collect baseline participant characteristics.

Before starting the multiweek training, we assessed gait, the endurance capacity of the calf muscles, ankle joint stiffness, muscle force characteristics and morphology, and motor unit firing properties. During the intervention, participants came to the lab almost every week for the first 7 weeks, and every two weeks thereafter. At each visit, we also took time to review and assess their exercise technique to ensure they were still performing it correctly and that the stimulus remained challenging and effective.

The data presented in this paper were recorded at following three time points, week 0 (baseline), week 6, and week 12.

### Eccentric training intervention

Participants completed a 12-week eccentric training program, with sessions performed on alternating days. Each session comprised four sets of 8-12 controlled eccentric contractions per leg, with 2 minutes of rest between sets. The primary exercise was a controlled heel drop, in which participants slowly lowered one heel into a full calf stretch and used the opposite leg to return to the tiptoe position. Each set was performed twice: once with the knee extended and once with a slight bend, to target different regions of the triceps Surae. From week 6, donkey calf raises were introduced, performed with the hips flexed and knees extended. Both straight- and bent-knee variations were included. Progressive overload was applied by adding 2.5 kg to a backpack once participants could complete 12 repetitions without fatigue. Training was avoided on the day prior to laboratory visits to minimise acute fatigue.

### 3-Dimensional Ultrasound

Calf muscle morphology was assessed using 3D ultrasound imaging as previously described by Weide et al., (2017). A 5 cm linear probe (Acuson S2000, 14L5 Probe, 4-15 MHz) was used in B-mode to capture images from the right leg. Participants were positioned on a Biodex bench (Biodex Medical Systems, Inc., New York) at 0° hip and knee flexion, with the hip and upper leg secured to prevent movement. Longitudinal ultrasound sweeps were performed from the medial border to the lateral end of the muscle, with 0.5 cm overlaps. The probe’s position and orientation were tracked via a motion capture system (Polaris Spectra Camera, Northern Digital Inc.) and synchronized with an external trigger. Data acquisition was performed using fcal software (PLUS Toolkit, Version 2.8.0. Perk Lab, Queen’s University, Kingston, ON, Canada). Calibration and 3D reconstruction were carried out using a custom MATLAB script (Weide et al., 2017) (The MathWorks, Inc. MATLAB R2024a. Natick, MA, USA).

To ensure consistency in muscle loading, a handheld dynamometer was used to determine the ankle angle corresponding to externally applied torques of 0 Nm and 4 Nm. The foot was then positioned accordingly for 3D measurements; in this study, we present data recorded at 4 Nm.

The 3D volume was analysed using ITK-SNAP (version 4.2.0, Yushkevich et al.,, 2006) as previously described by Weide et al.,, (2017).). Shortly, the muscle’s origin and distal end were localized in the mid-longitudinal plane (see for details Weide et al. 2017 The mid-longitudinal plane was divided into three equal-length length portions: proximal, mid, and distal regions, for fascicle length and pennation angle measurements. Muscle volume was determined through semi-automatic segmentation of anatomical cross-sections using ITK-SNAP, and PCSA was calculated by dividing muscle volume by fascicle length obtained, as well as pennation angles using a custom MATLAB script.

### Footplate angle-torque and footplate angle-fascicle length relationships

Participants were positioned prone on a Biodex dynamometer with the right lower leg secured to the footplate. Using an Acuson S2000 Ultrasound System with an 18L6 probe (HELX Evolution, Siemens Medical Solutions USA, Inc.), the midpoint of the gastrocnemius medialis muscle belly was identified. The probe was oriented to capture a true mid-longitudinal plane of the fascicles, with both ends visible between the deep and superficial aponeuroses and positioned as close to perpendicular to the aponeurosis as possible. Muscle fascicle length and pennation angle were measured at rest and during contraction.

Participants performed isometric maximal voluntary contractions at fixed footplate angles from maximum plantarflexion to maximum dorsiflexion, with torque recorded every 10° across the range of motion. At each angle, passive torque was measured immediately before contraction in the absence of voluntary effort. During voluntary contractions, fascicle length and pennation angle were recorded synchronously with torque and footplate angle.

For torque analysis, the gravitational moment of the foot and footplate was determined from passive trials, as described by Andersen et al., (2010), and subtracted from the torque signal. Active torque was calculated by subtracting passive torque from total torque. Torque-angle relationships (active and passive) were fitted with third- and second-order polynomials, respectively. Fascicle length-angle relationships were also modelled using polynomial fitting in Matlab.

Ultrasound images were captured using the Epiphan Capture Tool (Version 3.27.14.7; Epiphan Systems Inc., Ottawa, ON, Canada) and analysed using TimTrack (van der Zee & Kuo, 2022; van der Zee T. *TimTrack: an automated algorithm for estimating geometric features from ultrasound images*. GitHub; 2025; Available from: https://github.com/timvanderzee/ultrasound-automated-algorithm). Graphs were generated and statistical analyses performed using GraphPad Prism (Version 10.4.2; GraphPad Software, San Diego, CA, USA).

## Results

### Functional adaptations

#### Footplate angle-torque relationship

Active and passive normalized torque, plotted as a function of footplate angle (angle relative to the tibia), remained unchanged over time due to eccentric training (Fig. 3A). Mean torque values between 50° and 100° were similar at baseline, week 6, and week 12. However, some subjects exhibited substantial changes in the shape and magnitude of their torque-angle curves, suggesting an increase in torque-generating capacity in individual cases (Supplementary Fig. 1).

**Figure 1.**
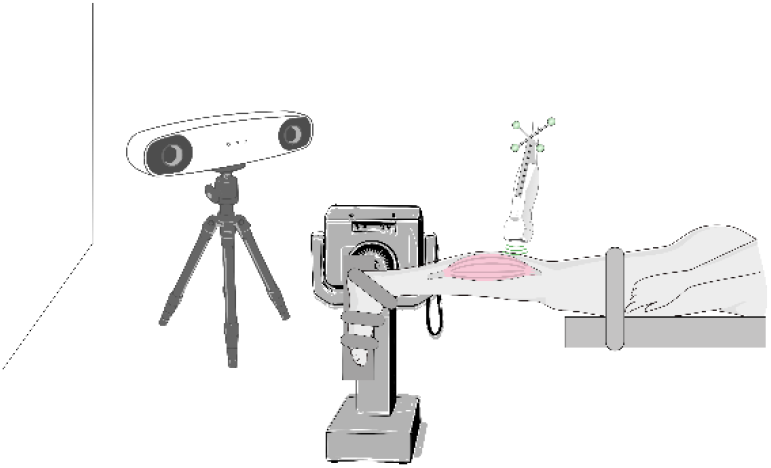
Experimental setup for musculoskeletal assessment combining 3D ultrasound (3DUS), optical tracking, and isometric testing on a dynamometer. Participants lie prone on a Biodex dynamometer bench with the hip and knee extended (0° flexion); the leg is positioned so that the knee extends just beyond the bench edge. The ankle angle is set to the position producing 4 Nm of passive plantarflexion torque, determined beforehand with a handheld dynamometer. At this angle, the foot is secured to the dynamometer footplate, and the leg is strapped to the bench to minimize movement. A linear-array transducer acquires B-mode images of the calf muscles while its position is tracked in real time by an optical system, enabling reconstruction of 3DUS volumes of the participant’s calf muscles under standardized mechanical loading.

**Figure 2.**
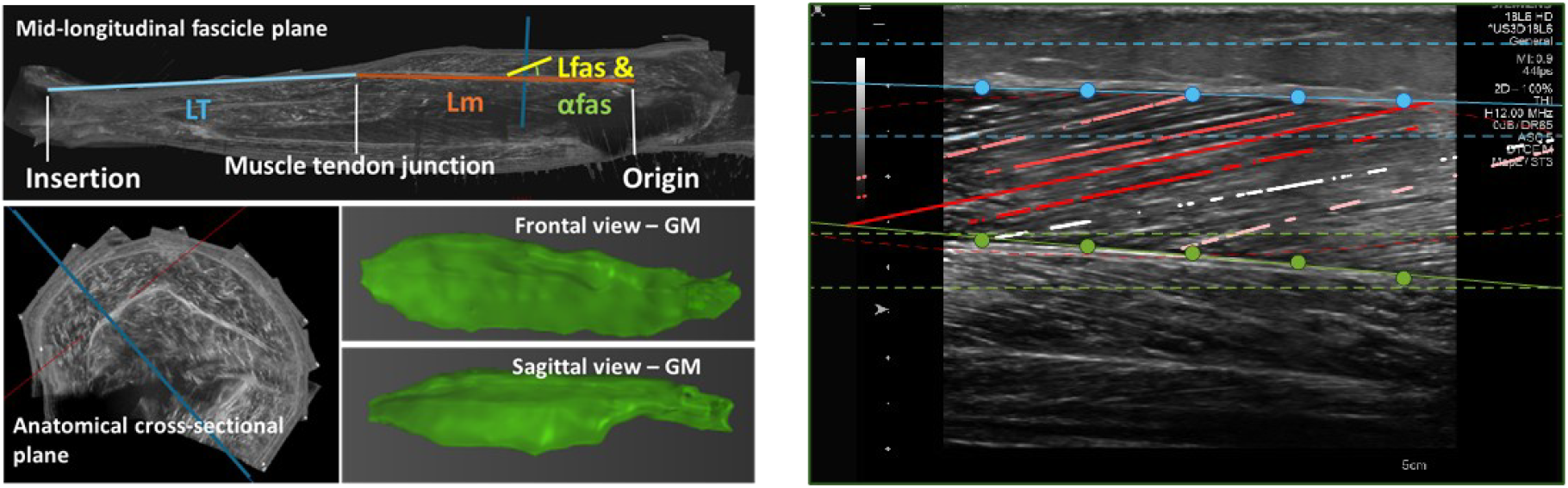
3D Reconstructions of the medial gastrocnemius muscle across a 12-week eccentric training intervention. **A**. Representative 3D ultrasound (3DUS) reconstruction of the medial gastrocnemius from a single participant, illustrating the acquisition and reconstruction procedure described in the text. **B**. Example fascicle-length and pennation angle measurement in the medial gastrocnemius using the *TimTrack* algorithm script at a fixed dynamometer footplate ankle angle while performing an isometric contraction.

**Figure 3.**
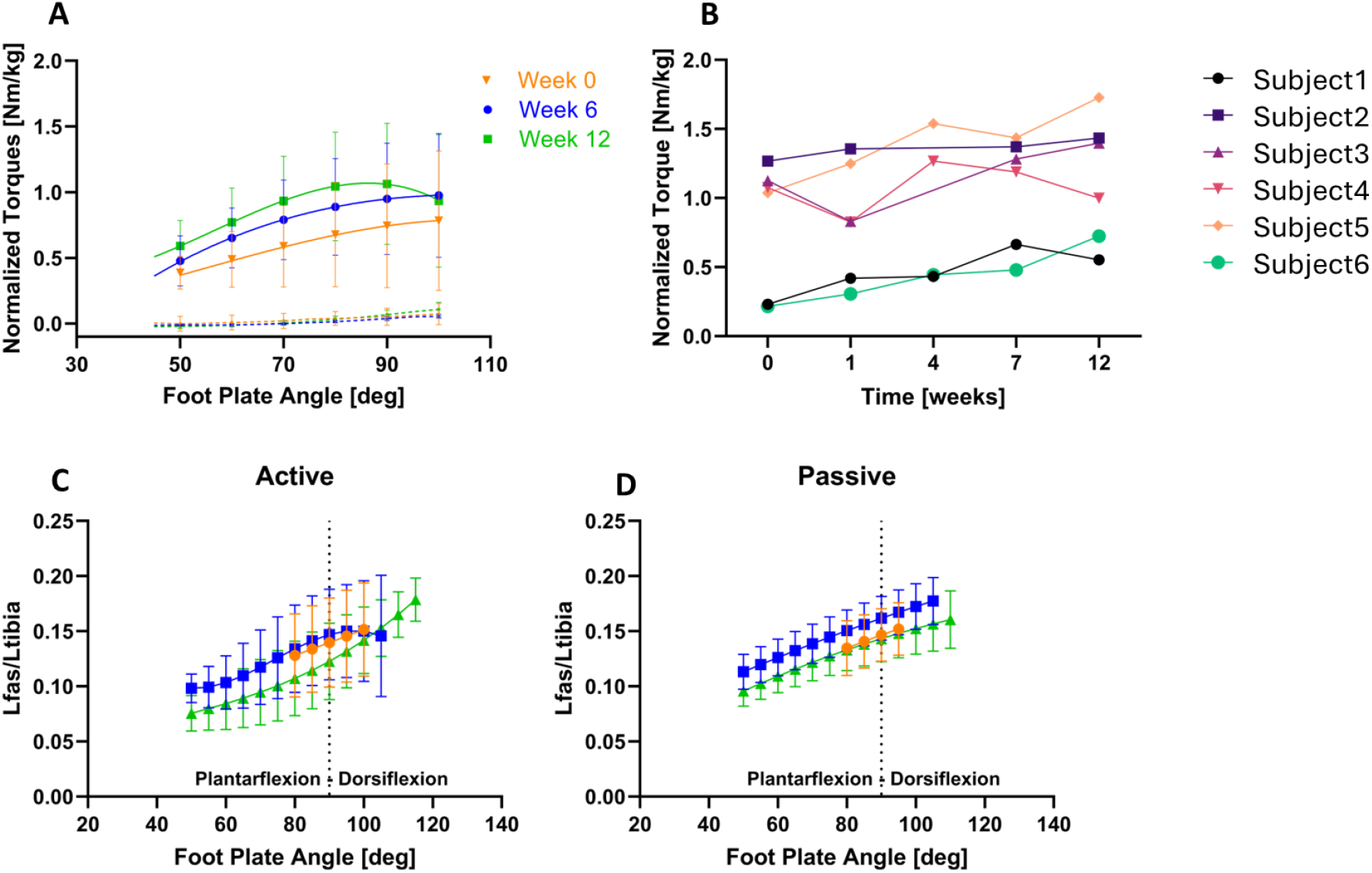
Longitudinal assessment of normalized torque-angle and fascicle-length relationships at baseline (week 0), mid-eccentric calf muscle training (week 6), and post-intervention (week 12). **A**. Active and passive footplate angle-torque relationship Shows torque (normalized to body weight) produced actively (during MVC) and passively across a range of footplate angles. Consistent with prior findings in triceps Surae, torque increases nonlinearly with angle due to changes in fascicle geometry. **B**. Time course of normalized MVC torque at 90° footplate angle Normalized torque measured during MVC at a fixed footplate angle of 90° across training weeks 0, 1, 4, 7, and 12. This panel captures strength adaptation over the intervention period. The graph shows a progressive increase in torque over time, with clear improvements from pre-to post-training, indicating enhanced force-generating capacity as a result of the training intervention. **C**. Passive footplate angle-fascicle length (Lfas) relationship. **D**. Active footplate angle-fascicle length relationship. ^*^ Shows p < 0.05. Results are provided as mean ± SD.

#### Peak force at 90° footplate angle

Normalized peak torque measured at a 90° footplate angle increased over time as a result of eccentric training (p = 0.01; Fig. 3B). Post hoc comparisons, corrected using the Bonferroni method for multiple comparisons, showed a 38.0% increase in torque of between baseline and week 12 (p = 0.01). These results indicate that maximal voluntary torque-generating capacity significantly improves after 7 weeks of eccentric training.

#### Footplate angle-fascicle length relationship

The relationship between footplate angle and fascicle length, measured at the mid muscle region both during active and passive conditions, remained similar following eccentric training at both week 6 and week 12 compared to baseline (Fig. 3C and 3D). Individual curves are presented in supplementary figure 2.

### Morphological adaptations, measured at 4Nm passive torque

#### Muscle belly and tendon lengths

The mean muscle belly and tendon lengths normalized to tibia length, as well as the normalized muscle-tendon complex length measured at 4Nm passive torque, remained similar following eccentric training at both week 6 and week 12 compared to baseline (Fig. 4A and 4B). However, Figure 4 highlights notable inter-individual variability: some subjects exhibited increased muscle belly length, while others showed a decrease. A similar pattern was observed for tendon length. Overall, changes in total muscle-tendon complex length varied among individuals, with some showing lengthening of the tendon, and others a reduction in the length of the entire complex.

**Figure 4.**
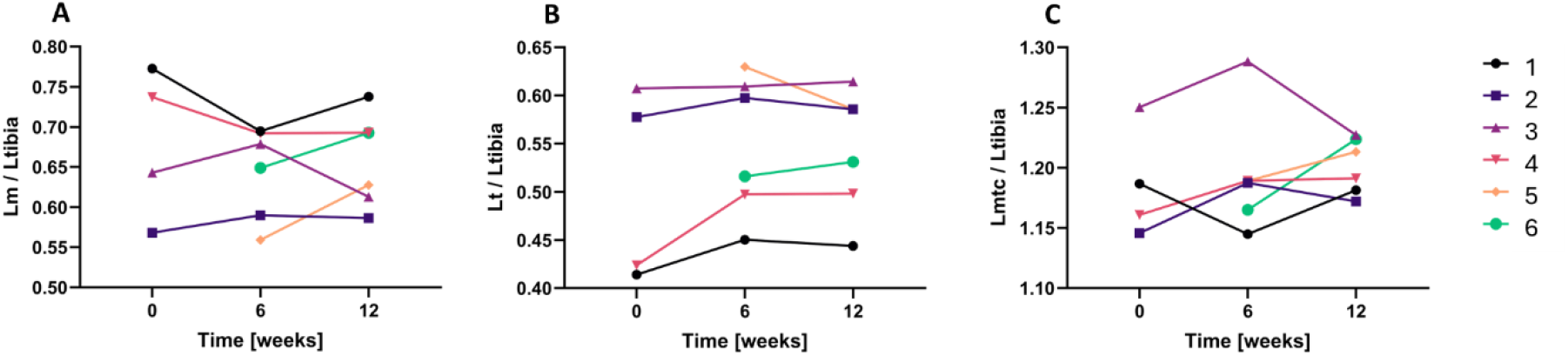
Longitudinal assessment of muscle-tendon complex morphology at baseline (week 0), mid-eccentric calf muscle training (week 6), and post-intervention (week 12). **A**. Normalized muscle length (Lm). **B**. Normalized tendon length (Lt). **C**. Normalized muscle-tendon complex length (Lmtc).

#### Muscle volume, measured at 4Nm passive torque

The muscle volume was similar between baseline, week 6, and week 12 (Fig. 5a). Muscle volume varied among individuals, with some showing slight increase of the volume, and others a reduction in the volume.

**Figure 5.**
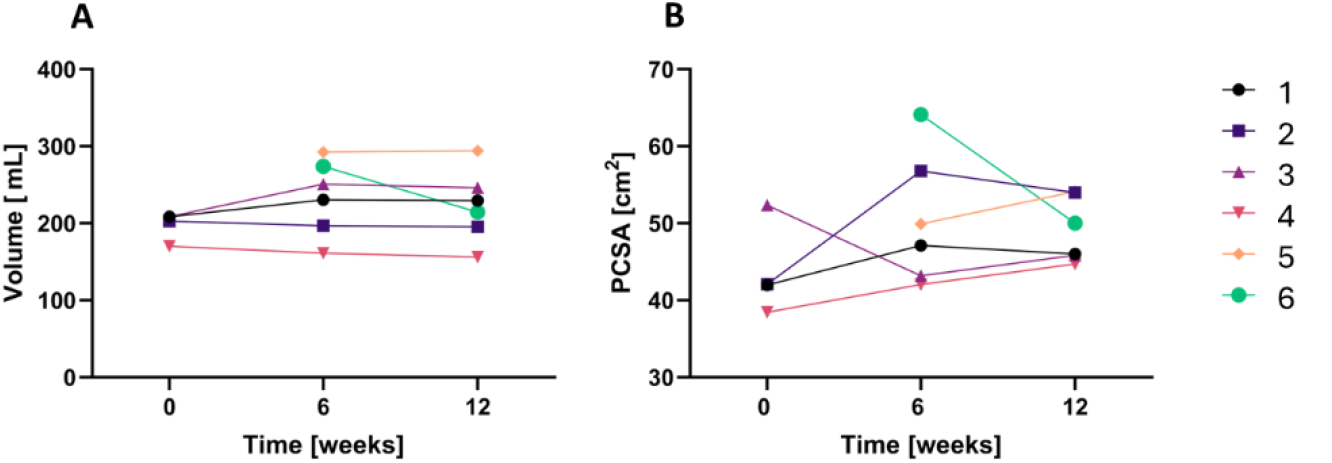
Longitudinal assessment of muscle volume and physiological cross-sectional area (PCSA) of the gastrocnemius medialis measured at a passive torque of 4 Nm at baseline (week 0), mid-eccentric calf muscle training (week 6), and post-intervention (week 12). **A**. Muscle volume of the gastrocnemius medialis measured using 3DUS. **B**. PCSA, computed as muscle volume divided by fascicle length measured in the mid-muscle region at the same passive torque.

#### Physiological cross-sectional area, measured at 4Nm passive torque

The PCSA was similar between baseline, week 6, and week 12 (Fig. 5B). The PCSA varied among individuals, with some showing slight increase of the area, and others a reduction in the area.

*Regional Muscle Adaptations: Fascicle Length and Pennation Angle at 4 Nm Passive Torque* Normalized mean fascicle lengths (relative to tibia length), measured at 4 Nm passive torque in the proximal, mid, and distal regions of the muscle, showed no changes following eccentric training at week 6 or week 12 compared to baseline (Fig. 6A). In the mid and distal region (Figs 6B and 6C), taken separately, the measured normalized fascicle length showed also similar values between baseline, week 6, and week 12. Similarly, pennation angles, which reflect the individual fascicle length patterns, also remained unchanged over time by trainig. These findings suggest that the training had overall no significant effect on fascicle length or pennation angle across muscle regions. Rather, results show inter-individual differences in adaptation.

**Figure 6.**
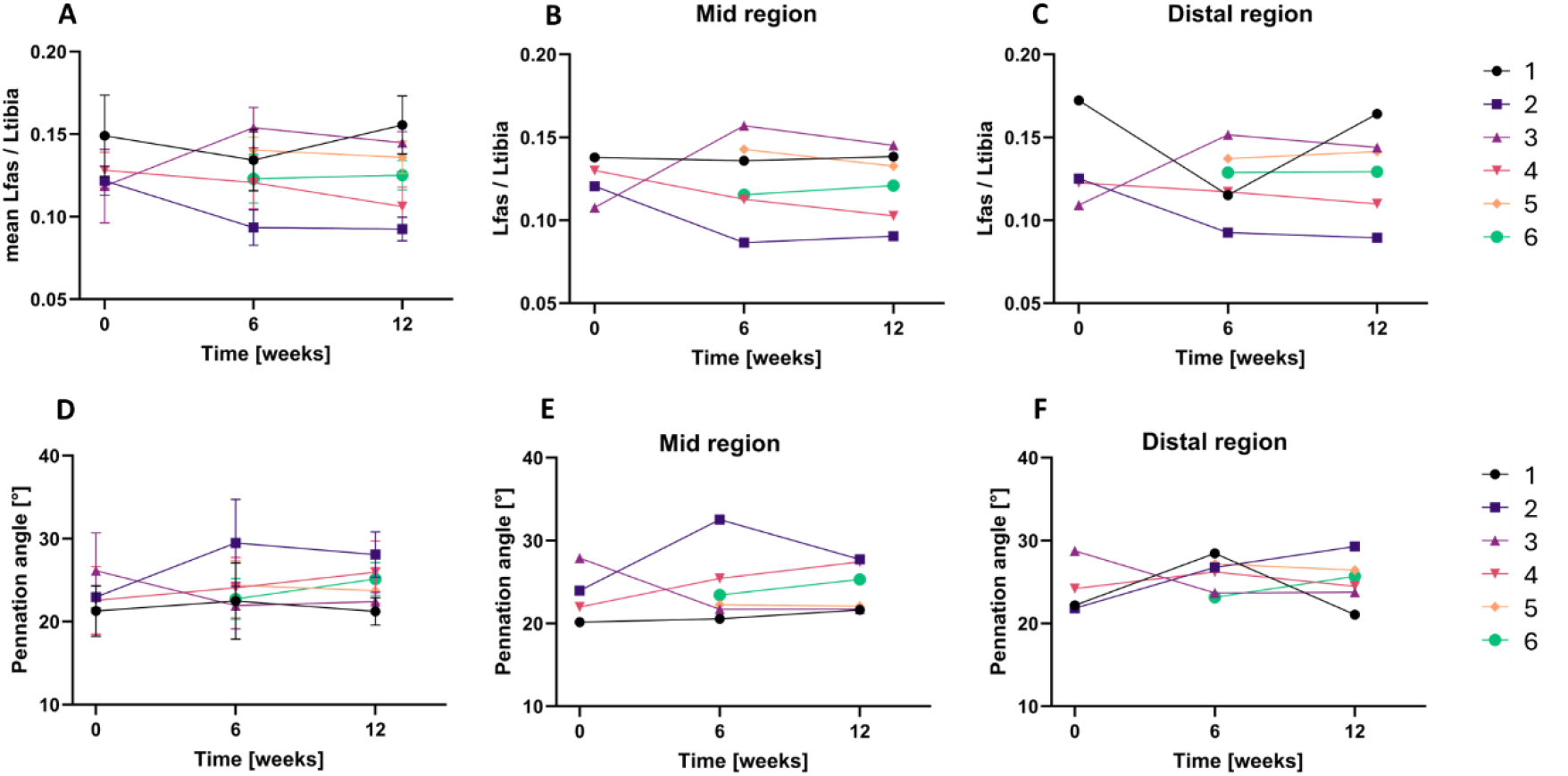
Longitudinal assessment of fascicle length (Lfas) and pennation angle (PA) of the medial gastrocnemius at baseline (week 0), mid-intervention (week 6), and post-intervention (week 12). **A.** Mean Lfas across the proximal, mid, and distal regions, normalized to tibia length. **B**. Mid-region Lfas normalized to tibia length. **C**. Distal-region Lfas normalized to tibia length. **D**. PA across the three regions. **E**. PA in the mid region. **F**. PA in the distal region.

### Individual muscle tendon complex adaptation profiles

#### A good responder

Subject 3 was considered a relatively good responder to the eccentric training intervention. Maximal torque in the footplate angle-torque relationship increased by 133%, and the optimal footplate angle for torque production shifted by **-**5.3%.

Gastrocnemius medialis volume measured at 4Nm passive torque increased by 18.1%, whereas PCSA decreased by 12.0%. Muscle length measured at 4Nm passive torque decreased by 4.7%, tendon length increased slightly by 1.2%, and total muscle–tendon complex length (normalized to tibia length) decreased by 1.8%.

At a passive force of 4Nm, fascicle lengths increased by 22.2%, while pennation angles decreased by 14.2%. The passive footplate angle-fascicle length relationship shifted right upwards, while the active relationship shifted left downward, indicating that fascicles operated at longer lengths during passive loading but shorter lengths at the same ankle angle during active contractions.

#### Less responsive participant

Subject 2 was considered a less responsive participant to the eccentric training intervention. The footplate angle-torque relationship showed no increase in maximal torque. At a passive force of 4 Nm, normalized gastrocnemius fascicle length decreased by 25%, while PCSA increased by 28% and volume decreased by 3.7%.

Gastrocnemius muscle length increased by 3.5%, tendon length increased by 1.7%, and total muscle-tendon complex length (normalized to tibia length) increased by 2.3%. For the footplate angle-fascicle length relationships, during the 6 and 12 weeks after training, the passive condition showed fascicles becoming shorter at smaller ankle angles compared to week 0. The active fascicle length-angle relationship shifted to the right, indicating that the same fascicle lengths occurred at higher ankle angles, and that the operating range extended to higher angles compared to baseline.

## Discussion

This study investigated temporal and regional adaptations in the medial gastrocnemius structure and function during a 12-week eccentric training program. In contrast to previously published findings reporting substantial increases in fascicle length and reductions in pennation angle after eccentric training (Duclay et al., 2009; Simpson et al., 2017; Geremia et al., 2019; Franchi et al., 2014; Raj et al., 2017; Bizet et al., 2025; Andrews et al., 2024; Pincheira et al., 2021), and clear hypertrophic responses in muscle volume or PCSA (Geremia et al.,, 2019; Andrews et al., 2024; Pincheira et al., 2021), group-level analyses here revealed no significant changes in fascicle length, pennation angle, muscle volume, or PCSA at either six or twelve weeks. Maximal voluntary torque at a 90° footplate angle increased moderately (38%), but other measures remained unchanged. Differences from prior reports likely reflect a combination of modest sample size, high inter-individual variance, and region-specific architectural remodelling. When variability is large relative to the effect size, mean values are insensitive even when several participants improve. Small but real changes, particularly in fascicle length or pennation angle, may also be obscured when responders and non-responders are pooled. Given this variability, examining individual adaptation profiles provides a clearer understanding of how the muscle responds to eccentric loading over time.

Subject 3 showed a pattern of adaptation typical for eccentric training, with large torque gains, longer fascicles, and smaller pennation angles-changes often linked to adding sarcomeres in series or increasing sarcomere length (Franchi et al., 2017; Pincheira et al., 2021). Muscle volume increased while PCSA decreased, suggesting longitudinal growth. The passive fascicle length-angle curve shifted upward, indicating longer fascicles under passive load, while the active curve shifted downward, suggesting increased tendon compliance and greater tendon length change during contraction. These changes coincided with a shift in optimal torque angle towards a more plantarflexed position.

In contrast, Subject 2 showed no improvement in the footplate angle torque relationship, though torque at 90° increased 11.1%. At baseline, this participant already had one of the highest normalized torque values in the group, as well as a distinct muscle-tendon architecture: a short muscle belly and a long tendon, resulting in one of the shortest fascicle lengths and highest pennation angles in the cohort. Like the other participants, he was a physically active, healthy individual, but not competing at a high level or as a professional athlete. Fascicle length at a standard passive load decreased further (-25%), while PCSA increased and volume decreased slightly, pointing to radial rather than longitudinal growth. Passive fascicles became shorter at smaller ankle angles, and the active fascicle length-angle curve shifted to the right, extending the operating range to higher angles. This pattern may reflect different tendon or muscle stiffness changes, which could limit translation of structural changes into torque gains (Reeves et al., 2003). The high starting torque, short fascicles, and long tendon may have limited the potential for large functional improvements. In a performance context, individuals with such a high baseline, similar to trained or non-amateur athletes, might still view even a small torque gain as meaningful, although such changes may not appear substantial in group-level data. Architectural changes in this participant were also not uniform along the muscle: fascicle length and pennation angle changes were more pronounced in the mid and distal regions than proximally, suggesting region-specific adaptation. Such patterns may arise from non-uniform strain distribution along the muscle-tendon unit, as shown in animal models (Kruse et al., 2021), reinforcing the idea that eccentric training induces both subject-specific and region-specific remodelling.

The contrast between subject 2 and subject 3 illustrates how baseline architecture, initial performance level, and tendon-muscle length ratios can shape the direction and magnitude of adaptation. Even under identical training protocols, adaptation can vary greatly between individuals in both magnitude and pattern. Some participants experience substantial gains in strength, muscle size, or architecture, while others exhibit minimal changes or adapt through different structural pathways, such as increases in fascicle length rather than PCSA. Such variability likely reflects differences in baseline muscle architecture and neural control, genetic predisposition, training compliance, or subtle differences in regional loading strategies, as well as broader inter-individual differences in responsiveness to training (Folland & Williams, 2007; Crouzier et al. 2018; Noone et al. 2024; Rong et al. 2025). Animal studies suggest that the muscle length or region under load critically determines the type and extent of adaptation, indicating that regional variation in stimulus (e.g., local strain or loading angle) could drive non-uniform architectural remodelling (Kruse et al., 2021). Understanding these factors is essential for interpreting training outcomes and for developing more targeted, individualized interventions.

Because of this wide range of individual responses, group-level averages can mask meaningful individual responses, a phenomenon well-documented in resistance training studies (Balshaw et al., 2017; Vellers et al., 2018). In our cohort, approximately half of the participants exceeded the smallest detectable change in torque-generating capacity or architectural measures, while others remained within measurement error. Because our outcomes reflect interrelated aspects of the same muscle, adaptation should be interpreted holistically rather than by examining isolated metrics. For example, torque gains without corresponding structural changes may indicate neural adaptation (Aagaard et al., 2001), whereas increases in PCSA or fascicle length without force improvements may represent structural remodelling that has yet to translate into performance. This multidimensional perspective is essential for accurately identifying responders and non-responders and for developing personalized interventions in both clinical and athletic populations.

A key limitation of this study is the small sample size, which reduces statistical power and increases the likelihood that true individual changes are diluted in group-level analyses. This constraint, combined with the high variability in adaptation profiles, means that the absence of significant group effects should be interpreted cautiously. The inclusion of both male and female participants may have introduced additional variability. For example, in female participants (Subjects 2 and 4), clear fluctuations in MVC at 90° were observed (Fig. 3B). Similar variability has been noted in trained females, where menstrual cycle phase and hormone levels can influence performance and recovery in some individuals (De Martin Topranin et al 2023; Engseth et al., 2023; Taylor et al., 2024), although group-level effects are inconsistent. We did not monitor menstrual cycle phase or contraceptive use, which limits interpretation of these fluctuations. While the primary aim was to capture and describe individual adaptation profiles, these factors highlight the importance of considering both absolute and relative changes when interpreting training outcomes. Nevertheless, the detailed, region-specific, longitudinal data collected here enable the identification of responder and non-responder profiles, support exploration of underlying mechanisms, and can inform the design of more targeted training interventions. These findings provide a foundation for larger studies to better understand long-term muscle remodelling in response to eccentric loading.

Beyond sample size and variability, a methodological limitation is that we did not control for the strain induced in the target muscle across participants. Because baseline architecture differs, the same eccentric training may impose different strains, potentially explaining part of the observed variability in adaptation. Incorporating real-time strain estimation in future work would allow training protocols to be tailored more precisely, paving the way for interventions that can reveal how specific mechanical stimuli drive muscle adaptation.

## Conclusion

This study uniquely tracked the time-course and regional architecture of the medial gastrocnemius in response to 12 weeks of eccentric training. Contrary to expectations, group-level architecture and morphology remained unchanged, though moderate improvements in maximal voluntary exerted torque were observed in some individuals. The pronounced inter-individual and region-specific variability underscores the importance of personalized profiling in training response. Beyond describing variability, these findings point toward the potential value of tailoring exercise interventions to baseline muscle–tendon architecture and regional strain patterns. Such individualized approaches could maximize functional gains in both clinical rehabilitation and athletic performance, and future research should test whether adapting load placement or joint angle emphasis can systematically optimize architectural remodelling.

## Data Availability

All data produced in the present study are available upon reasonable request to the authors

## Authors contribution

CR and MS conceived and designed the study. CR performed the experiments and collected the data. CR analysed the data and prepared the figures. MS and RJ assisted with data interpretation and critical revisions of the manuscript. CR drafted the manuscript. All authors contributed to the editing of the final version and approved the submitted manuscript.

## Conflicts of interest

The authors report no conflicts of interest in relation to the content of this manuscript.

The study protocol was reviewed and approved by the Ethics Committee of Twente (Enschede, The Netherlands) in accordance with the Declaration of Helsinki. All participants provided written informed consent prior to participation.

## Figures

**Supplementary Figure 1.**
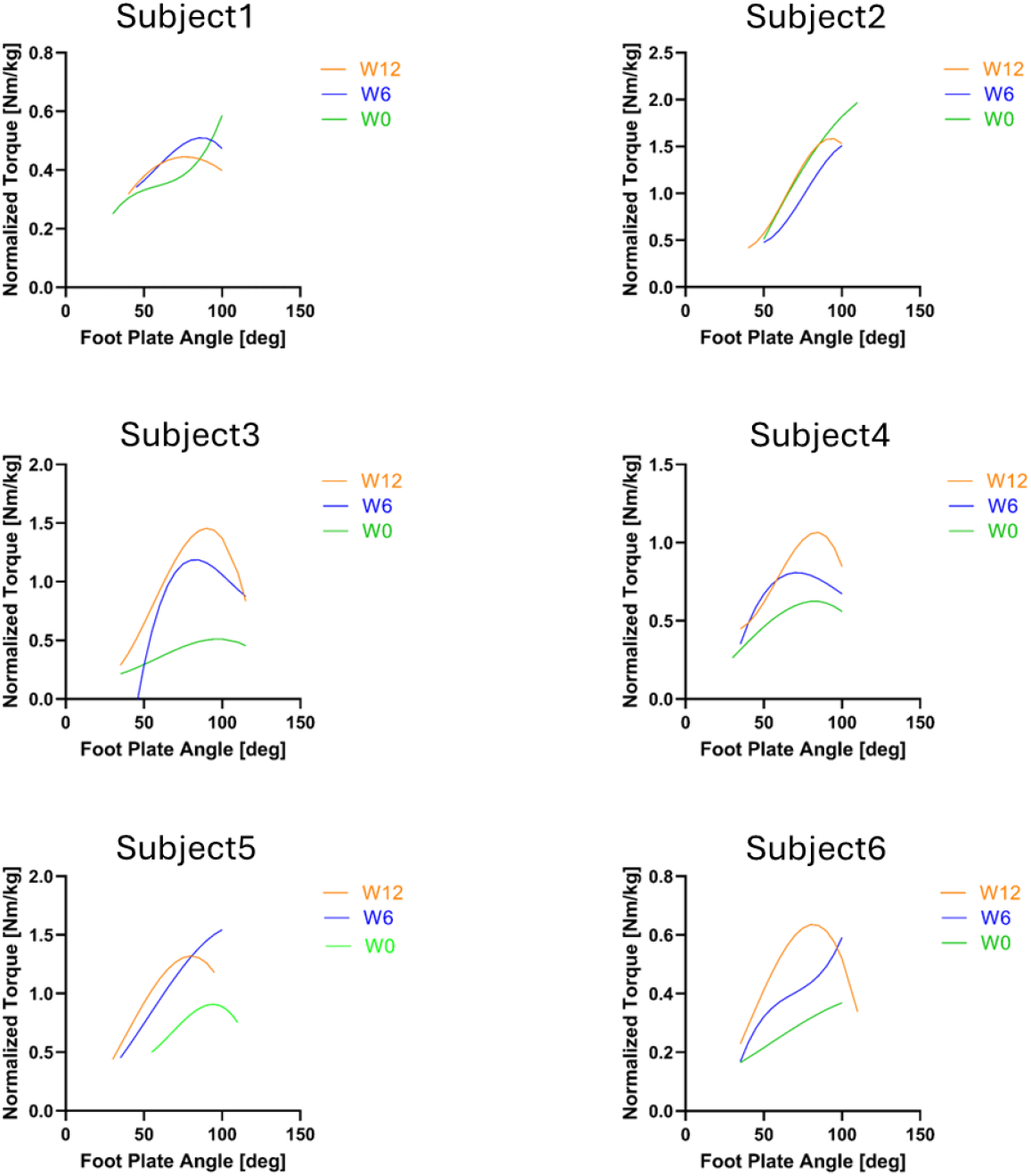
Individual longitudinal assessment footplate angle-torque relationships at baseline (week 0), mid-eccentric calf muscle training (week 6), and post-intervention (week 12). Individual active footplate angle-torque relationship.

**Supplementary Figure 2.**
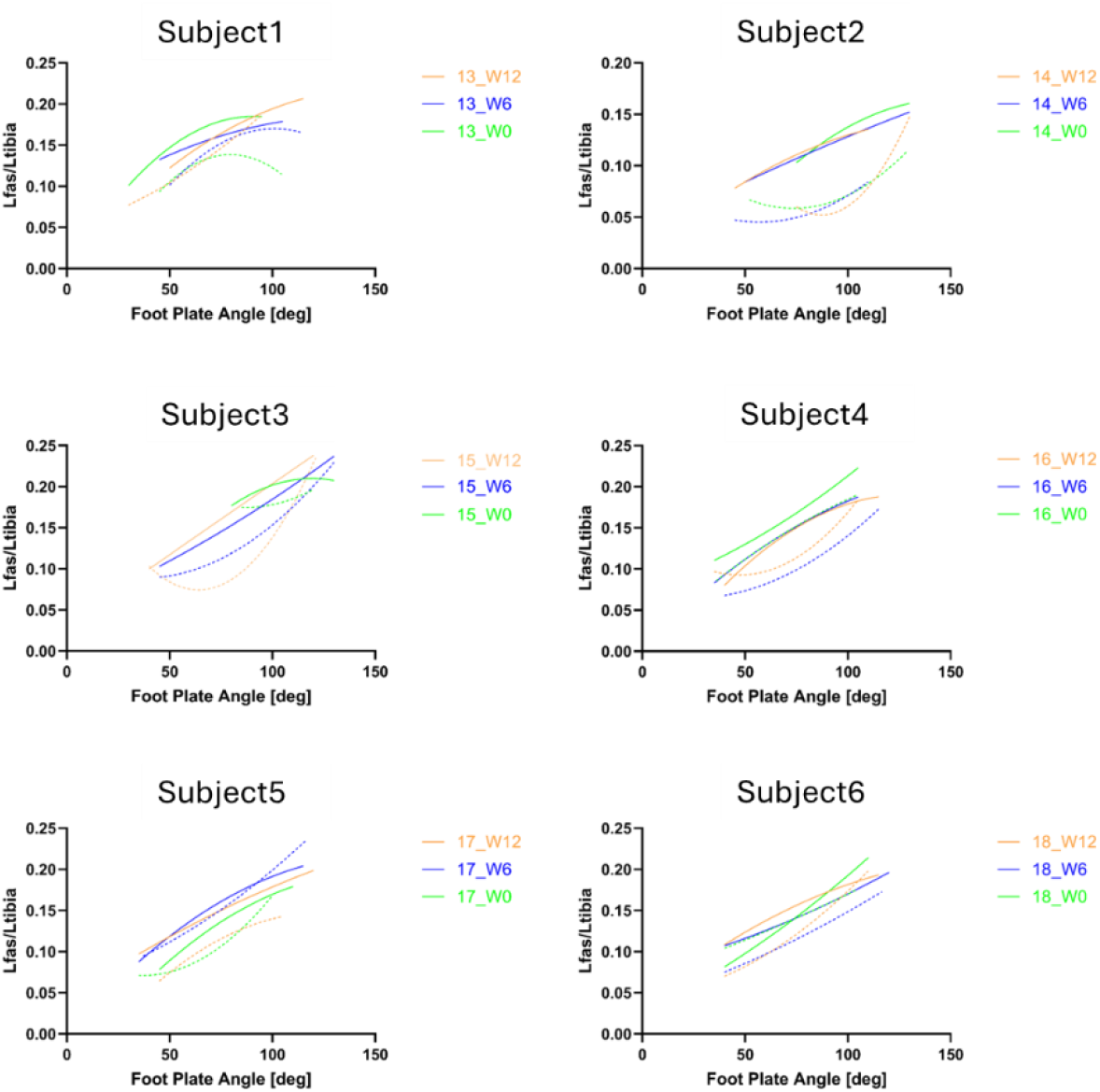
Individual longitudinal assessment footplate angle-fascicle length relationships at baseline (week 0), mid-eccentric calf muscle training (week 6), and post-intervention (week 12). Individual active and passive footplate angle-fascicle length (Lfas) relationship. Continuous lines represent the passive muscle fascicle length, while dashed lines represent the active length.

